# The effect of sepsis recognition on telemedicine use and bundle completion in rural emergency department sepsis treatment

**DOI:** 10.1101/2023.08.09.23293892

**Authors:** Anna M. Kaldjian, J. Priyanka Vakkalanka, Uche Okoro, Cole Wymore, Karisa K. Harland, Kalyn Campbell, Morgan B. Swanson, Brian M. Fuller, Brett Faine, Anne Zepeski, Edith A. Parker, Luke Mack, Amanda Bell, Katie DeJong, Kelli Wallace, Keith Mueller, Elizabeth Chrischilles, Christopher R. Carpenter, Michael P. Jones, Marcia M. Ward, Nicholas M. Mohr

**Author notes:** **Corresponding Author:** Nicholas M. Mohr, MD, MS, University of Iowa Carver College of Medicine 200 Hawkins Drive, Iowa City, IA 52242. **Institution Where the Work was Performed:** University of Iowa Carver College of Medicine, Iowa City, Iowa.

## Abstract

**Purpose:** Provider-to-provider emergency department telehealth (tele-ED) has been proposed to improve rural sepsis care. The objective of this study was to measure the association between sepsis documentation and tele-ED use, treatment guideline adherence, and mortality.

**Materials and Methods:** This analysis was a multicenter (n=23) cohort study of sepsis patients treated in rural emergency departments (EDs) that participated in a tele-ED network between August 2016 and June 2019. The primary exposure was whether sepsis was recognized in the local ED, and the primary outcome was rural tele-ED use, with secondary outcomes of time to tele-ED use, 3-hour guideline adherence, and in-hospital mortality.

**Results:** Data from 1,146 rural sepsis patients were included, 315 (27%) had tele-ED used, and 415 (36%) had sepsis recognized in the rural ED. Sepsis recognition was not independently associated with higher rates of tele-ED use (adjusted odds ratio [aOR] 1.23, 95% CI 0.90–1.67). Sepsis recognition was associated with earlier tele-ED activation (adjusted hazard ratio [aHR] 1.69, 95% CI 1.34-2.13), lower 3-hour guideline adherence (aOR 0.73, 95% CI: 0.55-0.97), and lower in-hospital mortality (aOR 0.72, 95% CI: 0.54-0.97).

**Conclusions:** Sepsis recognition in rural EDs participating in a tele-ED network was not associated with tele-ED use.

## Introduction

Sepsis contributes to 850,000 emergency department (ED) visits annually in the United States (U.S.).^1^ Of those with community-onset sepsis, mortality is 13%, making sepsis a leading cause of death in U.S. hospitals and a significant public health problem.^2, 3^ Sepsis also exhibits a significant volume-outcome relationship, with hospitals that have lower sepsis volumes having higher in-hospital sepsis mortality than those with high volumes.^4, 5^

Vague and varied symptoms at presentation make sepsis diagnosis challenging. Up to one-third of patients present without classic symptoms, and older and immunocompromised patients are particularly at risk for delayed recognition—possibly affecting early treatment and clinical outcomes.^6–8^ Variability in sepsis presentations has prompted the use of standardized tools to improve recognition, but universal recognition remains elusive.^9^

The Surviving Sepsis Campaign (SSC) publishes guidelines that recommend sepsis treatments, and timely high-quality sepsis care has been shown to improve clinical outcomes.^10, 11^ Telehealth has been proposed as one way to mitigate sepsis disparities in low-volume hospitals, bringing tertiary care expertise into low-volume EDs.^12^ Telehealth has been used in EDs for acute conditions such as ischemic stroke, myocardial infarction, and trauma, and it may impact care in sepsis.^12–18^

The objective of this study was to determine whether sepsis recognition in the ED is associated with ED-based telehealth (tele-ED) use, increased SSC bundle adherence, and decreased mortality. We performed a secondary analysis of the TELEmedicine as a Virtual Intervention for Sepsis in Emergency Departments (TELEVISED) study to identify (1) the proportion of rural sepsis patients recognized in local ED documentation, (2) predictors of local sepsis recognition, (3) the association between sepsis recognition, tele-ED use, and tele-ED timing, (4) association between sepsis recognition and SSC bundle adherence, and (5) the association between sepsis recognition and in-hospital mortality. We hypothesized that sepsis recognition would be associated with increased tele-ED use, higher SSC bundle adherence, and lower in-hospital mortality.

## Methods

### Study Design and Setting

This analysis is a cohort study conducted as a secondary analysis of the TELEVISED study, which was a retrospective, multicenter (n=23) study designed to measure the association between tele-ED use and clinical outcomes conducted in rural EDs between August 1, 2016 and June 30, 2019. Participating hospitals were rural as defined by the Federal Office of Rural Health Policy and took part in a single provider-to-provider tele-ED network.^19^ Detailed methods of the study and characteristics of included sites have been reported elsewhere, and the results of the main study have been previously published.^20, 21^ This study was approved by local institutional review boards under waiver of informed consent, and it is reported in accordance with the Strengthening Observational Studies in Epidemiology (STROBE) statement.^22^

### Selection of Patients

An electronic health record (EHR) data query was used to identify all adults (age≥18 years) diagnosed with sepsis in participating rural EDs during the study period. All of the following four inclusion criteria were required to meet our operational definition of ED sepsis as we have used previously (to account for explicit sepsis diagnosis codes in the ED being insensitive): (1) inpatient discharge diagnosis of sepsis by *International Classification of Diseases, 10^th^ edition, Clinical Modification* (ICD-10-CM) codes^20^, (2) infection suspected in the ED based on documentation in the ED clinical note (EHR), (3) organ failure in the ED (defined as a Sequential Organ Failure Assessment Score [SOFA] of 2 or greater or a change of at least 2 in patients with chronic disease), and (4) at least 2 systemic inflammatory response syndrome (SIRS) criteria in the ED.^23, 24^ Notably, patients could meet this criteria without sepsis being the ED diagnosis—even if formal sepsis diagnostic criteria were met.^23, 25^ We excluded any patient with no local diagnoses clearly documented in the EHR, as these medical records were likely incomplete. Data were abstracted by trained data abstractors using a structured data collection instrument according to the methods outlined by Kaji, et al.^26^

### Interventions

All EDs participated in a single hub-and-spoke tele-ED network and used a standard nurse-directed screening protocol. The local screening protocol was positive if (1) the local nurse “suspected infection” and (2) met 2 of the following criteria: temperature <96.8°F or >100.9°F, pulse >90 beats per minute, systolic blood pressure <90 mmHg, mean arterial blood pressure <65 mmHg, oxygen saturation <90%, respiratory rate >20 breaths/minute, lactate >2 mmol/L, white blood cell count <4,000 cells/μL or >12,000 cells/ μL. Local patients were screened by a triage nurse, and positive screens were reported to the local treating clinician. Tele-ED use was encouraged for patients screening positive, and we described the full screening protocol previously.^27^

The tele-ED network was an on-demand monthly subscription-based provider-to-provider real-time consultation service based in Sioux Falls, SD that could be activated by local providers or nurses 24 hours per day by pressing a button on the wall of the patient treatment room. Tele-ED consultation was provided by a board-certified emergency physician and an experienced ED nurse through a high-definition video connection. These consultants had access to the local EHR and were able to review records, place orders, provide real-time on-camera guidance, and arrange interhospital transfer when necessary.

### Exposures and Outcomes

Our primary exposure was whether sepsis was recognized in the rural ED (determined by EHR documentation), and our primary outcome was tele-ED use. Secondary outcomes were timing of tele-ED activation, SSC 3-hour bundle adherence (i.e., measure lactate, draw blood cultures before antibiotics, administer broad-spectrum antibiotics, and administer 30 mL/kg crystalloid fluid bolus if lactate is >4 mmol/L or systolic blood pressure is <90 mmHg), and in-hospital mortality.^10^

### Determination of Sepsis Recognition

Sepsis recognition was determined locally in the index ED. To determine whether sepsis was recognized locally, we conducted free-text analysis of ED diagnoses by two independent reviewers, who were trained using sample cases. All recorded diagnoses from the ED “impression” (not including differential diagnoses documented in the medical decision-making) were reviewed and classified into one of four mutually exclusive groups: (1) sepsis, (2) infection but no diagnosis of sepsis, (3) signs or symptoms that could be consistent with sepsis but without diagnosis of sepsis or infection, and (4) other unrelated diagnoses only. In this classification, sepsis included sepsis, septic shock, severe sepsis, or any other form of systemic response to infection (including SIRS as a diagnosis), but not including documentation of localized infection alone (e.g., septic arthritis). For example, a patient with an ED impression of “pneumonia” would be classified as “infection;” a patient with an ED impression of “hypoxia, hypotension, and acute kidney injury” would be diagnosed as “signs or symptoms consistent with sepsis;” and a diagnosis of “atrial fibrillation with rapid ventricular response” would be diagnosed as “other unrelated diagnoses only.” Notably, patients were included in the overall study if local clinicians documented in their medical decision-making that they suspected infection (e.g., as a differential diagnosis), but cases were not classified as infection unless an infection was listed in the “impression” portion of the note. Disagreements between the two reviewers were resolved by a third reviewer.

### Definitions

Fever was defined as temperature >38.0°C and hypothermia was defined as temperature <36.0°C. Hypotension was defined as systolic blood pressure <90 mmHg. Elevated lactate was defined as lactate >2 mmol/L. Tachycardia was defined as heart rate >100 beats/minute.

“Time zero” for time-based analyses was defined as the time of the initial set of vital signs in the ED when patients first met SIRS criteria for bundle adherence metrics and the time of ED arrival for time-to-tele-ED activation.

### Statistical Analysis

We calculated the percentage of locally recognized sepsis cases as the percentage of total included patients with an ED diagnosis of sepsis. Then we measured the association between recognized sepsis and variables of interest, using univariate logistic regression. Variables of interest were selected *a priori* and included source of infection, fever, hypothermia, hypotension, tachycardia, elevated lactate, surgery during hospitalization, SOFA score, and ED provider type (advanced practice provider [APP, either solo provider or provider with on-site physician] vs. physician only).

We used multivariable generalized estimating equations (GEE) with a logit link to identify predictors of locally recognized sepsis using our *a priori* variables. We then removed variables based on statistical criteria using the Quasi-Likelihood Information Criterion (QIC) to achieve parsimony, we required tele-ED use to remain in the model, and we included hospital identifier as a random effect to account for clustering.^28^ All covariates were categorical, and we confirmed model goodness of fit.^29^ We screened for collinearity and interaction terms. Then we constructed another GEE model using similar methods to measure the association between sepsis recognition (exposure) and 3-hour SSC bundle adherence (outcome) and a separate model to measure the association between sepsis recognition (exposure) and in-hospital mortality (outcome).

Among patients who had tele-ED used, we compared time-to-tele-ED activation using Kaplan-Meier curves and the log-rank test to identify whether patients with recognized sepsis had faster tele-ED activation. Data for this analysis were right censored if patients left the emergency department or died. Then, we used a Cox proportional hazards model adjusted for the same covariates to measure the association (adjusted hazard ratio [aHR]) between sepsis recognition (exposure) and time-to-tele-ED (outcome). We assessed the proportional hazards assumption of the adjusted model using a global test based on the Schoenfeld residuals.

Analyses were conducted as complete case analysis without imputation, but missing values on the variables of interest were rare. All statistical procedures were conducted using two-tailed tests, p<0.05 was considered statistically significant, and analyses were conducted using SAS v 9.4 (SAS Institute, Cary, NC).

## Results

### Baseline Characteristics

We included 1146 patients (**Figure 1**), and 315 (27%) had tele-ED used. Patients had a median age of 72 years (interquartile range [IQR] 62 – 82), and 517 (45%) were female. The most common source of infection was pneumonia (n=619, 54%), and the median ED sequential organ failure assessment (SOFA) score was 4 (IQR 3 – 5). A total of 347 (30%) patients were transferred to another hospital, and 111 (10%) patients died in the hospital. Baseline characteristics are detailed in **Table 1**.

**Figure 1.**
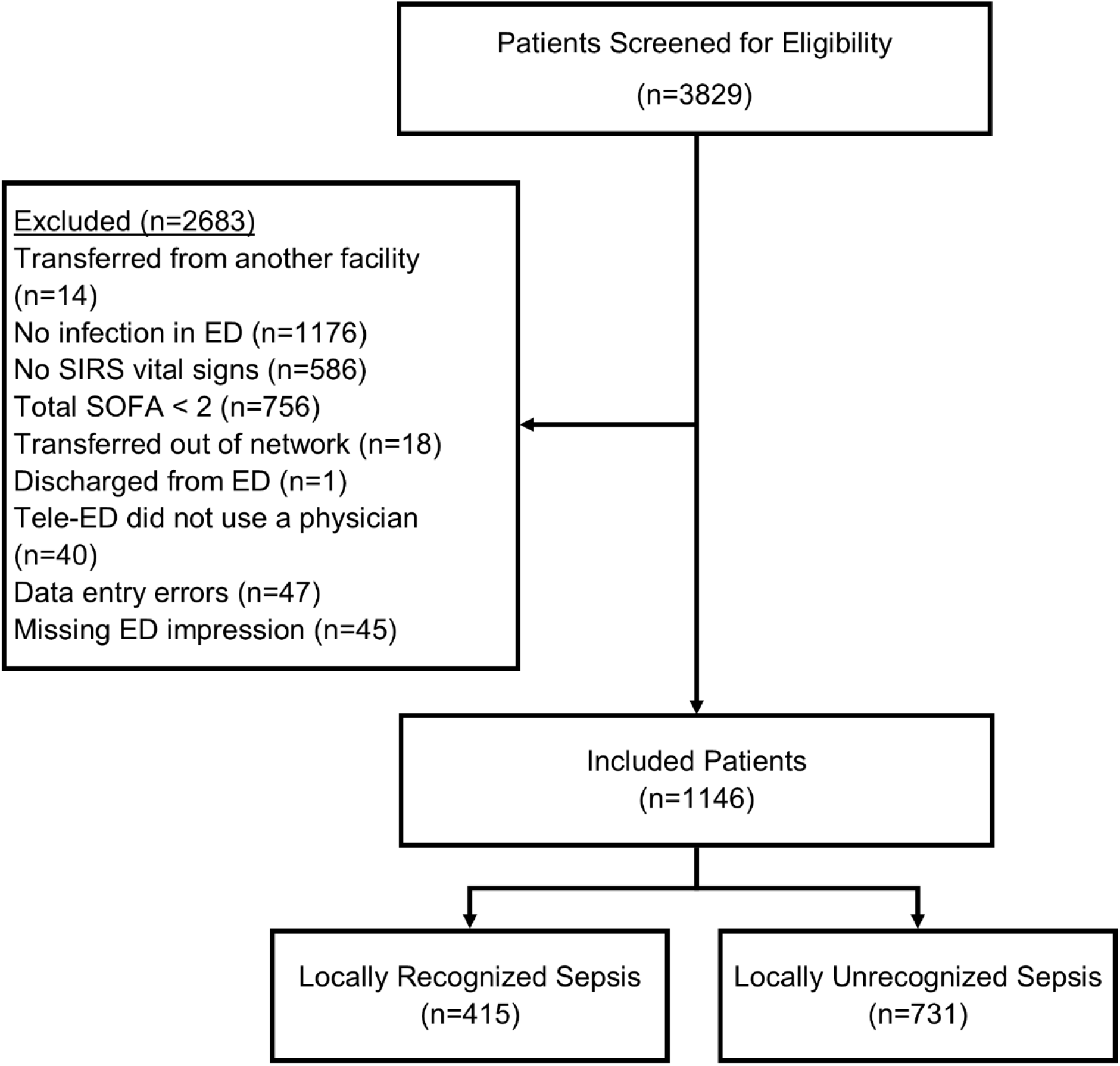
Selection of participants.

**Table 1.** Characteristics of participants. IQR, interquartile range; kg, kilograms; m, meters, 95% CI, 95% confidence interval; SOFA, Sequential Organ Failure Assessment; APP, advanced practice provider; COPD, chronic obstructive pulmonary disease

|  | All Cases<br>(n=1146)<br>n (%) | No Local<br>Sepsis<br>Recognition<br>(n=731)<br>n (%) | Local<br>Sepsis<br>Recognition<br>(n=415)<br>n (%) | Difference<br>(95% CI) |
| --- | --- | --- | --- | --- |
| **DEMOGRAPHICS** |  |  |  |  |
| Age, years |  |  |  |  |
| <30 | 9 (0.8) | 5 (0.7) | 4 (1.0) | 0.3 (-1.4, 0.8) |
| ≥30 and <40 | 38 (3.3) | 26 (3.6) | 12 (2.9) | 0.8 (-1.4, 2.8) |
| ≥40 and <50 | 68 (5.9) | 43 (5.9) | 25 (6.0) | 0.1 (-3.0, 2.8) |
| ≥50 and <60 | 116 (10.1) | 64 (8.8) | 52 (12.5) | 3.7 (-0.1, 7.5) |
| ≥60 and <70 | 291 (25.4) | 178 (24.4) | 113 (27.2) | 2.8 (-2.5, 8.1) |
| ≥70 and <80 | 267 (23.3) | 174 (23.8) | 93 (22.4) | 1.4 (-3.7, 6.5) |
| ≥80 and <90 | 274 (23.9) | 185 (25.3) | 89 (21.5) | 3.8 (-1.3, 8.9) |
| ≥90 | 83 (7.2) | 56 (7.7) | 27 (6.5) | 1.2 (-1.9, 4.3) |
| Age, median (IQR) | 72.0 (62.4 – 82.1) | 73.6 (62.9 – 82.9) | 70.1 (61.4 – 80.8) | 3.5 (1.0, 6.1) |
| Female | 517 (45.1) | 332 (45.4) | 185 (44.6) | 0.8 (-5.2, 6.8) |
| Body mass index, kg/m<sup>2</sup> |  |  |  |  |
| <18.5 | 39 (3.6) | 28 (4.0) | 11 (2.9) | 1.1 (-1.1, 3.3) |
| ≥18.5 and < 25 | 283 (26.4) | 177 (25.5) | 106 (28.0) | 2.5 (-3.1, 8.1) |
| ≥25 and <30 | 290 (27.0) | 191 (27.5) | 99 (26.1) | 1.4 (-6.9, 4.1) |
| ≥30 and <35 | 198 (18.5) | 128 (18.4) | 70 (18.5) | 0.1 (-4.8, 5.0) |
| ≥35 | 263 (24.5) | 170 (24.5) | 93 (24.5) | 0 (-5.4, 5.4) |
| Latinx | 13 (1.2) | 6 (0.8) | 7 (1.8) | 1.0 (-0.5, 2.5) |
| Race |  |  |  |  |
| White | 1016 (89.4) | 652 (90.1) | 364 (88.4) | 1.7 (-2.1, 5.5) |
| African American | 8 (0.7) | 3 (0.4) | 5 (1.2) | 0.6 (-0.4, 2.0) |
| American Indian or Alaska Native | 92 (8.1) | 57 (7.9) | 35 (8.5) | 0.6 (-2.7, 3.9) |
| Other | 20 (1.8) | 12 (1.7) | 8 (1.9) | 0.2 (-1.4, 1.8) |
| **PATIENT CHARACTERISTICS** |  |  |  |  |
| Triage systolic blood pressure, mmHg, median (IQR) | 130 (109 – 149) | 131 (112 – 151) | 126 (104 – 147) | 5.0 (0.5, 9.5) |
| SOFA Score |  |  |  |  |
| 2–3 | 560 (48.9) | 372 (50.9) | 188 (45.3) | 5.6 (-0.4, 11.6) |
| 4–7 | 521 (45.5) | 332 (45.4) | 189 (45.5) | 0.1 (-5.9, 6.1) |
| ≥8 | 65 (5.7) | 27 (3.7) | 38 (9.2) | 5.5 (2.4, 8.6) |
| SOFA Score, median (IQR) | 4 (3 - 5) | 3 (3 – 5) | 4 (3 – 5) | 1 (0.7, 1.3) |
| Source of Infection |  |  |  |  |
| Respiratory | 619 (54.0) | 441 (60.3) | 178 (42.9) | 17.2 (11.5, 23.3) |
| Genitourinary | 209 (18.2) | 125 (17.1) | 84 (20.2) | 3.1 (-1.6, 7.8) |
| Abdominal | 69 (6.0) | 43 (5.9) | 26 (6.3) | 2.9 (-2.5, 3.3) |
| Device-Related | 2 (0.2) | 0 (0) | 2 (0.5) | 0.5 (-0.2, 1.2) |
| Skin/Soft Tissue | 109 (9.5) | 74 (10.1) | 35 (8.4) | 1.7 (-0.5, 1.8) |
| Meningitis | 13 (1.1) | 7 (1.0) | 6 (1.5) | 0.5 (-0.9, 1.9) |
| Unknown | 125 (10.9) | 41 (5.6) | 84 (20.2) | 14.6 (10.4, 18.8) |
| Surgery during hospital stay | 77 (6.7) | 43 (5.9) | 34 (8.2) | 2.3 (-0.8, 5.4) |
| Interhospital Transfer | 347 (30.3) | 196 (26.8) | 151 (36.4) | 9.6 (4.0, 15.2) |
| Length of hospital stay (days), median (IQR) | 4.9 (3.2 – 8.3) | 4.9 (3.2 – 8.0) | 5.0 (3.1 – 9.0) | 0.1 (-0.5, 0.6) |
| Provider Type |  |  |  |  |
| Physician | 975 (85.1) | 635 (86.9) | 340 (81.9) | 5.0 (0.6, 9.4) |
| APP with physician | 112 (9.8) | 56 (7.7) | 56 (13.5) | 5.8 (2.0, 9.6) |
| APP alone | 59 (5.2) | 40 (5.5) | 19 (4.6) | 0.9 (-3.5, 17.1) |
| Arrive by Ambulance | 561 (50.0) | 341 (47.4) | 220 (54.7) | 7.3 (-1.2, 13.4) |
| In-Hospital Mortality | 111 (9.9) | 64 (9.0) | 47 (11.5) | 2.5 (-1.2, 6.2) |
| **COMORBIDITIES** |  |  |  |  |
| COPD | 361 (31.5) | 259 (35.4) | 102 (24.6) | 10.8 (5.4, 16.2) |
| Cirrhosis | 30 (2.6) | 16 (2.2) | 14 (3.4) | 1.2 (-0.8, 3.2) |
| Solid organ transplant | 12 (1.1) | 7 (1.0) | 5 (1.2) | 0.2 (-1.1, 1.5) |
| Cancer | 339 (29.6) | 221 (30.2) | 118 (28.4) | 1.8 (-3.9, 7.1) |
| Diabetes | 426 (37.2) | 281 (38.4) | 145 (34.9) | 3.5 (-2.3, 9.3) |
| Chronic Dialysis | 45 (3.9) | 28 (3.8) | 17 (4.1) | 0.3 (-2.1, 2.7) |
| Asthma | 138 (12.0) | 93 (12.7) | 45 (10.8) | 1.9 (-1.9, 5.7) |
| Hypertension | 824 (71.9) | 531 (73.0) | 293 (70.6) | 2.4 (-3.0, 7.8) |

### Locally recognized sepsis

In our sample, 415 (36%) had sepsis recognition documented in the local ED. Other non-sepsis local diagnoses included infection (n=624, 54%); signs, symptoms, or findings consistent with sepsis (n=85, 7%); and an unrelated diagnosis only (n=22, 2%). Sepsis was more likely to be diagnosed in patients who presented with fever (41% vs. 33%, difference 8%, 95% CI 2–14%), hypotension (63% vs. 34%, difference 29%, 95% CI 19–39%), or higher SOFA scores (53% with SOFA 7 or greater vs. 34% with SOFA 2–6, difference 19%, 95% CI 9–29%). Sepsis recognition was higher in those treated by an APP (with or without onsite physician supervision) compared to those treated by a physician alone (44% vs. 35%, difference 9%, 95% CI 1–17%). Tele-ED was used more frequently in those with sepsis recognition compared with those without (45% vs. 23%, difference 13%, 95% CI 7–19%). In multivariable modeling, fever, hypotension, tachycardia, elevated lactate, and APP-directed care (both alone and with an onsite physician) remained predictive of increased odds of locally recognized sepsis, and pneumonia (compared with all other infection sources) was associated with decreased local recognition of sepsis. Tele-ED use was not associated with sepsis recognition in multivariable modeling (aOR 1.23, 95% CI 0.90–1.67, **Figure 2**).

**Figure 2.**
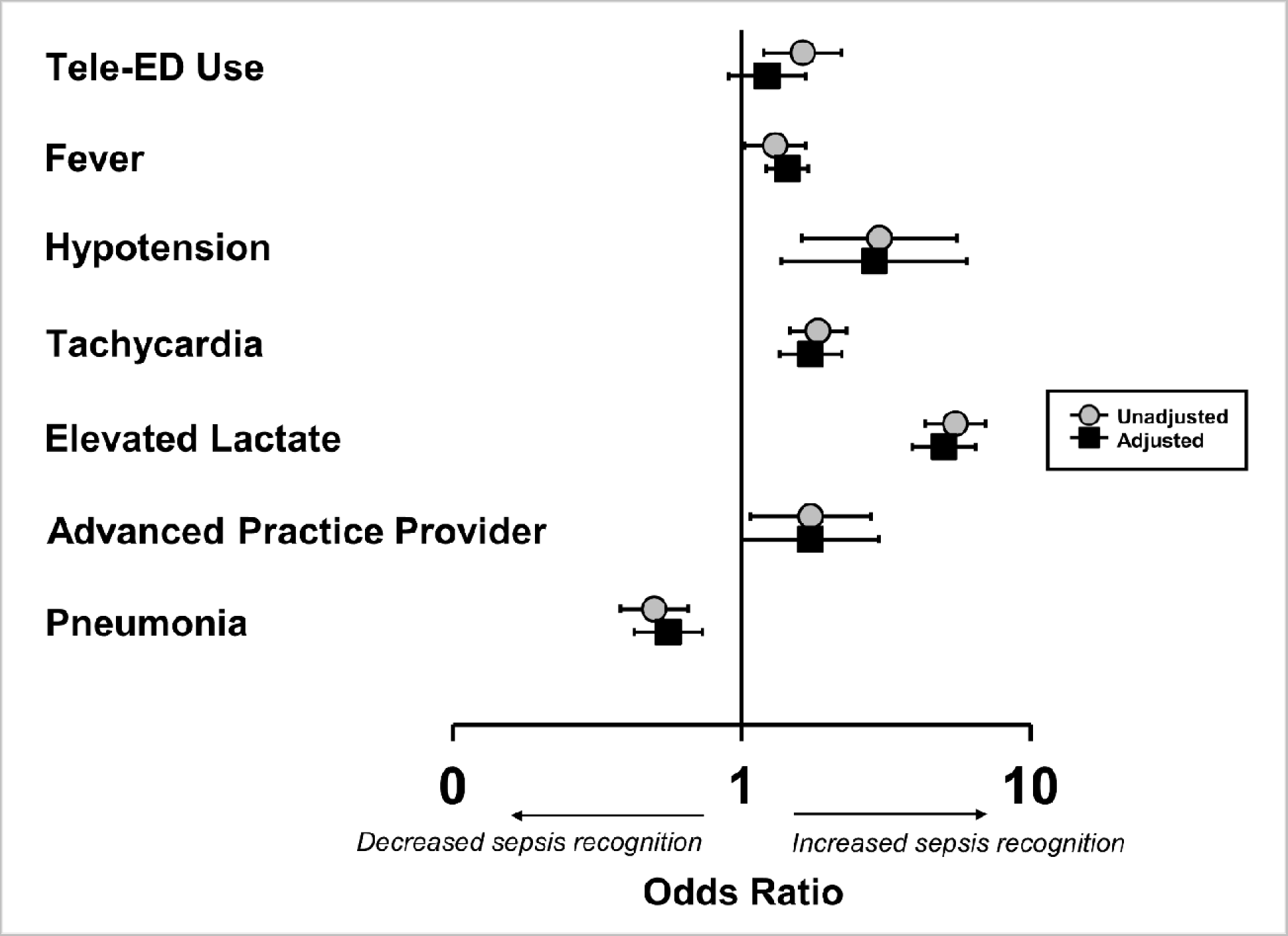
Factors predicting local recognition of sepsis. This forest plot depicts the association between signs and symptoms and local recognition of sepsis. Grey circles represent unadjusted values and black squares represent adjusted values. Variables used in statistical adjustment include all those variables shown in this forest plot. *ED, emergency department*

The hazard function associated with tele-ED activation was 64% earlier in those with recognized sepsis versus those without (unadjusted HR 1.64, 95% CI 1.31–2.06), and this difference persisted when adjusted for ED disposition and provider type (aHR 1.69, 95% CI 1.34–2.13, **Figure 3**).

**Figure 3.**
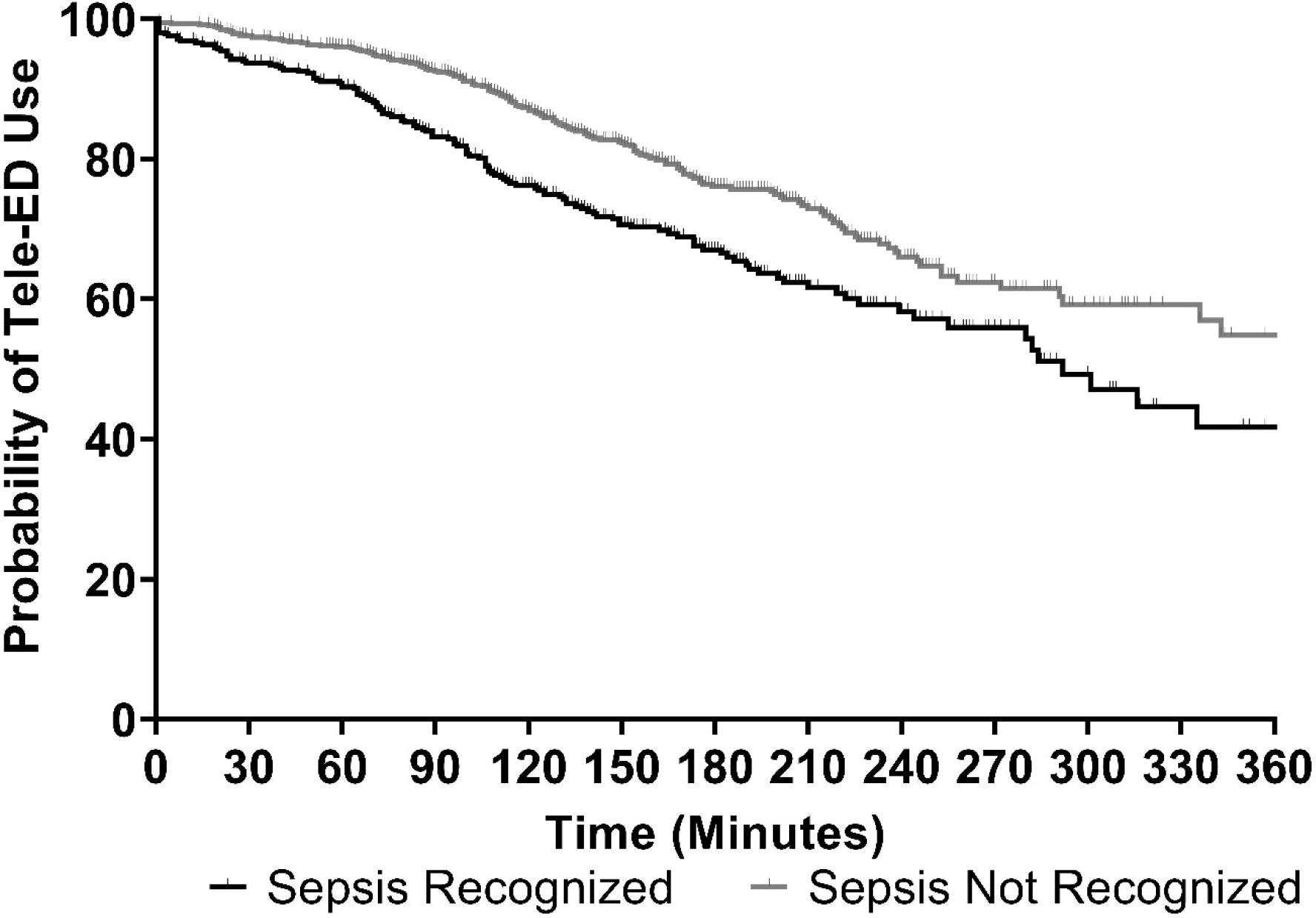
Time to tele-ED use, stratified by sepsis recognition. This figure depicts the Kaplan-Meier survival plot of time to tele-ED consultation. Data are right censored if patients left the ED or died. *ED, emergency department; APP, advanced practice provider*.

### SSC bundle adherence and in-hospital mortality

SSC 3-hr bundle adherence was similar between those with and without locally recognized sepsis in bivariate analyses (15% vs. 12%, difference 2.9%, 95%CI −1.3–7.1%). After covariate adjustment, sepsis recognition was associated with lower 3-hr bundle adherence (aOR 0.73, 95% CI 0.55–0.97). In-hospital mortality was similar in patients who had local sepsis recognition (11% vs. 9%, difference 2.5%, 95%CI −1.2–6.3%); however, after adjusting for covariates, in-hospital mortality was significantly lower in those with local sepsis recognition (aOR 0.72, 95% CI 0.54–0.97, **Figure 4**).

**Figure 4.**
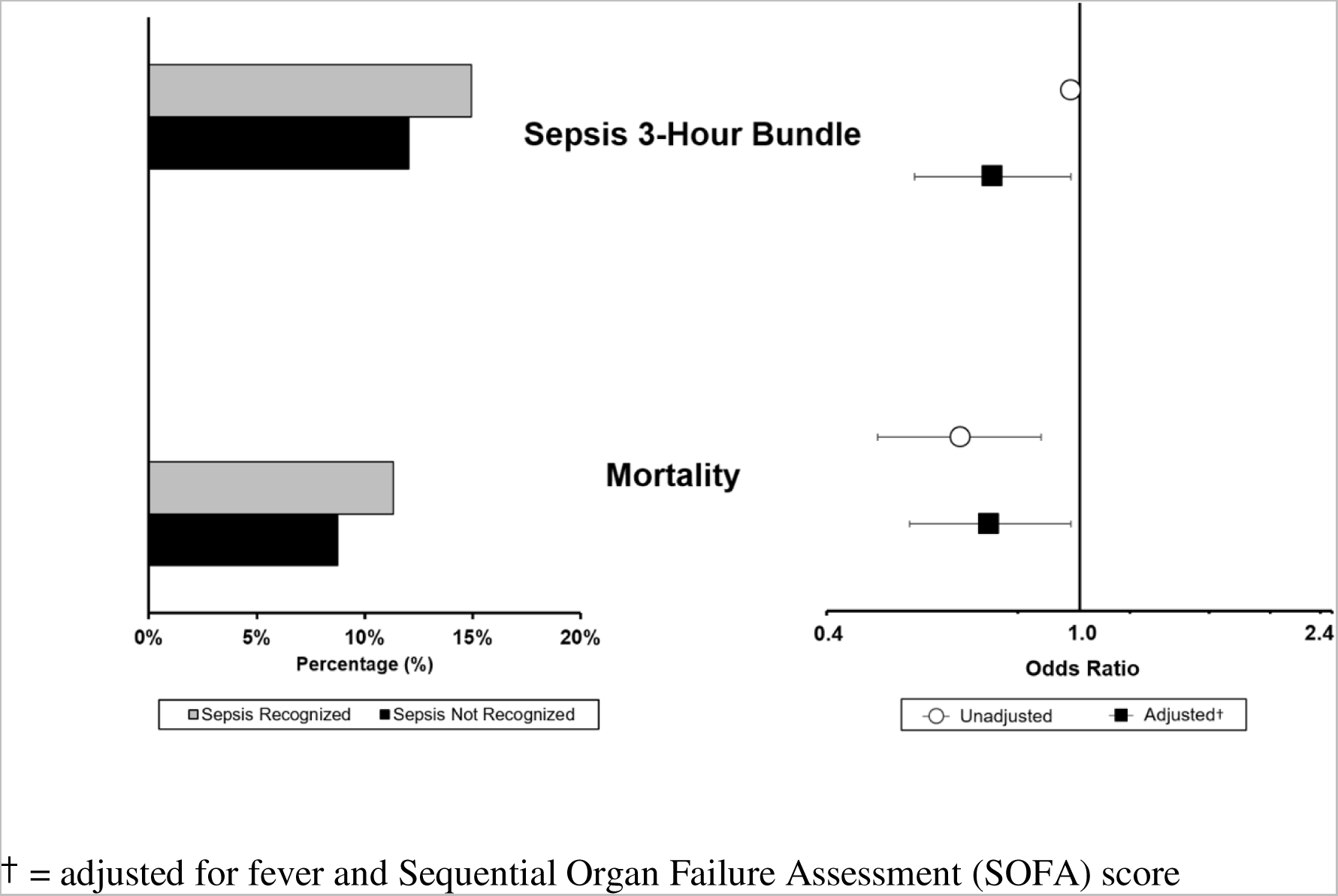
Surviving Sepsis Campaign (SSC) 3-hour bundle adherence and in-hospital mortality, stratified on sepsis recognition. This figure depicts the association between local sepsis recognition, SSC bundle adherence (3-hour), and in-hospital mortality. On the left panel, the raw adherence numbers are reported (gray bars represent recognized sepsis, and black bars represent unrecognized sepsis). On the right panel, the unadjusted and adjusted odds ratios of local sepsis recognition and each outcome are presented (log scale, white circles represent unadjusted analyses, and black squares represent adjusted analyses). Error bars represent 95% confidence intervals. Note that the point estimate and confidence interval for the unadjusted estimate for the 3-hour bundle is hidden by the symbol, but it is OR 0.97 (95% CI 0.95, 1.00).

## Discussion

In our analysis of a multicenter cohort study in rural EDs, we found that tele-ED use was more common in those recognized as having sepsis. However, adjusting for factors associated with sepsis recognition, tele-ED use was not independently associated with sepsis recognition. Furthermore, we observed that a minority of sepsis patients in participating rural hospitals were recognized as having sepsis—regardless of tele-ED use. In our prior report of the TELEVISED study, we showed that tele-ED use was not associated with improved clinical outcomes or SSC bundle adherence.^21^ The finding from this analysis is relevant, because under-recognition could be a key explanation for why our prior work did not show the differences in processes and outcomes we hypothesized we might observe.

Sepsis diagnosis—especially for patients with low illness severity—can be difficult, and under-recognition is an obstacle to appropriate and timely sepsis care.^7, 30^ In a cohort study of 654 ED sepsis patients, 37% presented with vague symptoms, and the absence of explicit, traditional symptoms of sepsis was associated with delays in antibiotic therapy and higher mortality.^6^ Others have associated misdiagnosis with delayed treatment and worse clinical outcomes.^31^ This finding parallels our results in that sepsis recognition was associated with decreased adjusted in-hospital mortality, but we additionally found that the failure to recognize sepsis was associated with a lower frequency of tele-ED use.^18^

Many patients in whom sepsis was not recognized were appropriately diagnosed as having pneumonia, respiratory failure, or kidney injury—all diagnoses that these patients clearly had. One of the reasons sepsis diagnoses may be missed is that sepsis may be a secondary syndrome diagnosed in someone who may appropriately have a primary diagnosis of infection.^32^ Biomarkers have been proposed as one strategy to improve recognition of the constellation of findings that suggest a host immune response and may define sepsis, and EHR-based algorithms and ICU-based telehealth screening have also been proposed.^33–35^

Provider-to-provider telehealth has been viewed as a major advantage for managing rare, life-threatening conditions in low-volume centers.^36^ However, our findings highlight one significant shortcoming of this approach—when tele-ED use is low, appropriate on-demand tele-ED activation requires accurate local diagnosis.^37^ Under-recognition of sepsis has been commonly reported.^31, 38, 39^ Similar patterns have emerged in stroke, myocardial infarction, and aortic dissection, where treatment paradigms are clear and under-recognition is associated with morbidity and mortality.^40–42^ Like other time-sensitive acute care conditions, sepsis treatment has been operationally protocolized, so tele-ED’s greatest potential to improve guideline adherence and outcomes may be from its ability to identify sepsis and initiate a predefined treatment bundle. The fact that remote diagnostic surveillance screening is not a feature of most current tele-ED programs is a significant concern, and it may be a call to action to implement EHR-based and telemetry-based strategies to incorporate into tele-ED networks screening technology for conditions not recognized by local providers. Informatics and machine learning-based tools continue to improve, and providing health system-level surveillance—even remotely—may be one way to amplify the effect of tele-consultation by remote experts.^43^

Our study has several limitations. First, we relied on EHR documentation to indicate recognition of sepsis or infection, and such documentation may miss cases accurately recognized (but not adequately documented) by the treating clinician. Lack of documentation in the medical record has been hypothesized to decrease case capture for the sepsis core measure, but as most of our sites do not voluntarily report sepsis quality measures^44^, we think this factor is not likely relevant in our sample. Second, our observational study cannot be used to imply causality, because we do not know whether tele-ED consultation is caused by sepsis recognition, diagnosis is caused by tele-ED consultation, or if clinical features that drive tele-ED consultation are coincidentally associated with accurate sepsis diagnosis. Finally, our multicenter study was conducted in a single tele-ED network that used common procedures at all sites, which may affect the generalizability of our results.

In conclusion, sepsis recognition in rural EDs participating in a tele-ED network was not independently associated with tele-ED use, but recognition was associated with decreased adjusted in-hospital mortality. Future work will seek to identify ways to improve rural sepsis recognition using automated methods and tele-ED surveillance, and to identify barriers to effective tele-ED use.

## Data Availability

All data produced in the present study are available upon reasonable request to the authors.

## ACKNOWLEDGEMENTS

The authors would like to acknowledge Sydney Krispin, MA, MPH for her editorial assistance in preparing this manuscript.

